# sSocial Determinants of Health and Cumulative Incidence of Mortality among U.S. Adults without Major Chronic Diseases

**DOI:** 10.1101/2024.03.16.24304395

**Authors:** Sophie E. Claudel, Ashish Verma

**Affiliations:** Department of Medicine, Boston Medical Center, Boston, MA, USA; Section of Nephrology, Department of Medicine, Boston University Chobanian and Avedisian School of Medicine, Boston, MA, USA

**Keywords:** social determinants of health, mortality, food security, income, education, NHANES, cumulative incidence

## Abstract

**Background:** Social determinants of health (SDOH) are widely known to contribute to poor health outcomes and premature mortality among individuals with prevalent diseases. Understanding the impact of SDOH on mortality among adults without major chronic diseases may inform public policy in the United States (U.S.).

**Methods:** We conducted a prospective observational study using the National Health and Nutrition Examination Survey data (1999-2018) among 11,413 adults without hypertension, diabetes, hyperlipidemia, severe obesity, chronic kidney disease, cardiovascular disease, chronic respiratory disease, cancer, liver disease, arthritis, hepatitis B or C, human immunodeficiency virus, or pregnancy. We calculated 15-year adjusted cumulative incidences of all-cause mortality by baseline SDOH and described the trends in the prevalence of adverse SDOH.

**Results:** The mean ±SD age was 34.9±11.2 years. Over a median follow-up of 10.3 years, 275 participants died. The prevalence of low educational attainment decreased over the study period from 19.8% to 12.1%, while the prevalence of food insecurity increased from 11.5% to 23.1%. The 15-year adjusted cumulative incidences of all-cause mortality were 5.6% (95% CI, 2.8-8.5), 5.2% (95%CI, 3.2-7.3), 4.9% (95%CI, 2.7-7.2), 4.0% (95%CI, 2.8-5.2) for participants who had < 100 % of the federal poverty level, below high school education, food insecurity, and were born in the U.S., respectively. In the final adjusted model, a 1-point increase in cumulative SDOH score was associated with 33% increased risk of all-cause mortality.

**Conclusions:** In this study of community-dwelling U.S. adults without major chronic diseases, we demonstrate a pronounced increase in all-cause mortality associated with adverse SDOH.

## Introduction

Countries where investment in social services exceeds direct medical expenditure demonstrate improved population health outcomes compared to those where the reverse is true, such as the United States (U.S.).^1^ The prevailing hypothesis behind these findings is that addressing ‘upstream’ causes of disease, or social determinants of health (SDOH), conveys more significant benefits than mitigating individual-level health risks.^2,3^ SDOH – including food insecurity, economic disadvantage, and low educational attainment – are widely known to contribute to poor health outcomes and premature mortality among individuals with prevalent diseases.^4–7^ SDOH are hypothesized to act, at least partially, via epigenetic, inflammatory, or hormonal pathways to impact health negatively.^4,8–10^

While SDOH has been shown to exacerbate numerous chronic diseases, relatively little is known about how SDOH impacts the prognosis of healthy adults. Understanding the role of adverse SDOH in determining risk for poor outcomes in states of apparent health may provide valuable insights into preventative health strategies upstream of the healthcare system. This study aimed to examine the adjusted cumulative incidence of mortality associated with adverse SDOH among U.S. adults without major chronic diseases and describe the prevalence of these SDOH between 1999-2018.

## Methods

### Study Population

We included adults aged 20 and older who participated in the National Health and Nutrition Examination Survey (NHANES) 1999-2018. NHANES is a biannual, cross-sectional survey of the non-institutionalized U.S. population that includes demographic, social, and medical surveys, in addition to laboratory and examination data.^11^ We excluded participants with diabetes (defined by hemoglobin A1c (HbA1c) ≥6.5% or use of hypoglycemic medications), hypertension (defined by systolic blood pressure ≥130 mmHg or diastolic blood pressure ≥80 mmHg, or use of antihypertensive medications), cardiovascular disease (including self-reported angina, heart disease, stroke, heart failure, or myocardial infarction), hyperlipidemia, estimated glomerular filtration rate (eGFR) <60 ml/min per 1.73m^2^, urine albumin to creatinine ratio ≥30 mg/g, arthritis, hepatitis B, hepatitis C, human immunodeficiency virus, chronic respiratory disease (asthma, emphysema, chronic obstructive pulmonary disease, or chronic bronchitis), liver disease of any kind, any sort of malignancy, those who received dialysis in the past 12 months, and those who were pregnant at the time of the exam. Details of chronic disease determination are described in **Table S1**. The final sample size was 11,413.

### Ascertainment of Social Determinants of Health (SDOH)

We included seven SDOH (income, education, employment, food security, nativity and acculturation, health insurance, and access to a routine source of healthcare) based on participants’ responses to NHANES questionnaires. Variable definitions are outlined in **Table S2**. Poverty status was defined as <100%, 100-300%, or ≥300% of the Federal Poverty Level based on the family poverty-to-income ratio (PIR). We categorized educational attainment as less than high school, high school graduate (or General Education Diploma), and above high school education. Occupational status was determined by response to the kind of work the participant did last week. If a respondent indicated they were not working due to being a student or retired, they were coded as ‘employed.’^5^ Food security was dichotomized based on responses to the U.S. Food Security Survey as food secure (indicating no affirmative responses) versus food insecure (marginal, low, or very low food security).^12^ Nativity and acculturation were determined based on self-reported country of birth and length of residence in the U.S. We included two variables related to access to healthcare, including lack of health insurance and a routine source of medical care (other than the emergency department).

### Ascertainment of Mortality

We linked the participants to the National Death Index to determine all-cause mortality. Deaths were ascertained through December 31, 2019.

### Covariates

We included the following covariates based on clinical and biological plausibility to affect the relationship between SDOH and mortality: age, sex, race or ethnicity (non-Hispanic White, non-Hispanic Black, Mexican American, other Hispanic, and other race), body mass index (BMI, kg/m^2^), total cholesterol (mg/dL), systolic blood pressure (mmHg), smoking status (never, former, current), heavy alcohol use (average of ≥4 drinks day among women or ≥5 drinks day among men over the past 12 months),^13^ current or former injection drug use, HbA1c (%), and eGFR (ml/min per 1.73m^2^). eGFR was calculated using the 2021 CKD-EPICr equation.^14^

### Imputation

We used hot deck multiple imputation (with simple random sampling with replacement), which accounts for complex survey procedures, due to missingness in the following variables (**Table S3**): serum creatinine (N=900), total cholesterol (N=876), systolic blood pressure (N=541), BMI (N=105), HbA1c (N=19), and smoking status (N=12).^15^ **Table S4** shows the participant characteristics prior to imputation for comparison.

### Statistical Analysis

We summarized descriptive statistics of the study population using weighted means (95% confidence interval (CI)) or weighted percentages, as appropriate. We calculated the weighted, age-adjusted prevalence of each SDOH and age-standardized the estimates to the 2010 U.S. Census. We created four pre-determined periods for analysis to minimize the effect of small sample sizes: 1999-2002, 2003-2008, 2009-2014, and 2015-2018. We used standard Wald tests from survey logistic regression models to obtain *p*-values for temporal trends, controlling for age.^16^

We calculated the 10- and 15-year adjusted cumulative incidences using confounder-adjusted survival curves employing the G-Formula method for covariate adjustment.^17^ We calculated absolute risk differences and the corresponding 95% CI at 15 years of follow-up. We plotted adjusted survival curves using the R software package “adjusted curves” using final, survey-weighted adjusted multivariable-adjusted Cox proportional hazard models.^17,18^ The adjustment strategy was based on covariates’ biological and clinical plausibility as potential confounders of the association between SDOH and mortality. Models were adjusted for age, sex, race or ethnicity, survey year, smoking status, systolic blood pressure, total cholesterol, HbA1c, BMI, eGFR, alcohol use, and injection drug use.

We additionally examined the relative risk of mortality associated with adverse SDOH using multivariable-adjusted survey-based Cox regression models. Model 1 was adjusted for age, sex, race or ethnicity, and survey year. Model 2 additionally adjusted for smoking status, systolic blood pressure, BMI, HbA1c, total cholesterol, and eGFR. Model 3 additionally adjusted for alcohol use and injection drug use.

All analyses accounted for the complex survey design of NHANES except for the cumulative incidence plot of the cumulative SDOH variable. Analyses were performed in SAS (version 9.4) or R software (version 3.5). A two-tailed p-value <0.05 was considered statistically significant. All data are publicly available at https://wwwn.cdc.gov/nchs/nhanes/default.aspx.

## Results

The mean (±SD) age was 34.9±11.2 years and 51.9% were female (**Table S5**). The median follow-up time was 10.3 years and 275 participants died. Individuals who self-identified as non-Hispanic Black and Hispanic appeared to have lower income, lower educational attainment, and greater food insecurity than non-Hispanic White participants (**Table S5**).

### Prevalence of Adverse Social Determinants of Health

Trends in the prevalence of adverse SDOH are shown in **Figure 1** and summarized in **Table S6**. The overall prevalence of poverty-to-income ratio ≤100% of the federal poverty level, below high school education, and food insecurity were 11.8% (95% CI 10.8-12.8), 15.5% (95% CI 14.1-17.0), and 17.5% (95% CI 16.2-18.9), respectively. There was a decrease in the proportion of adults with a below-high school education and those without health insurance between 1999-2018. The prevalence of food insecurity increased between 1999-2018, from 11.5% (95% CI 9.6-13.4) to 23.1 (95% CI 18.9-27.3). Poverty, employment, and access to a routine source of healthcare did not change over the study period.

**Figure 1.**
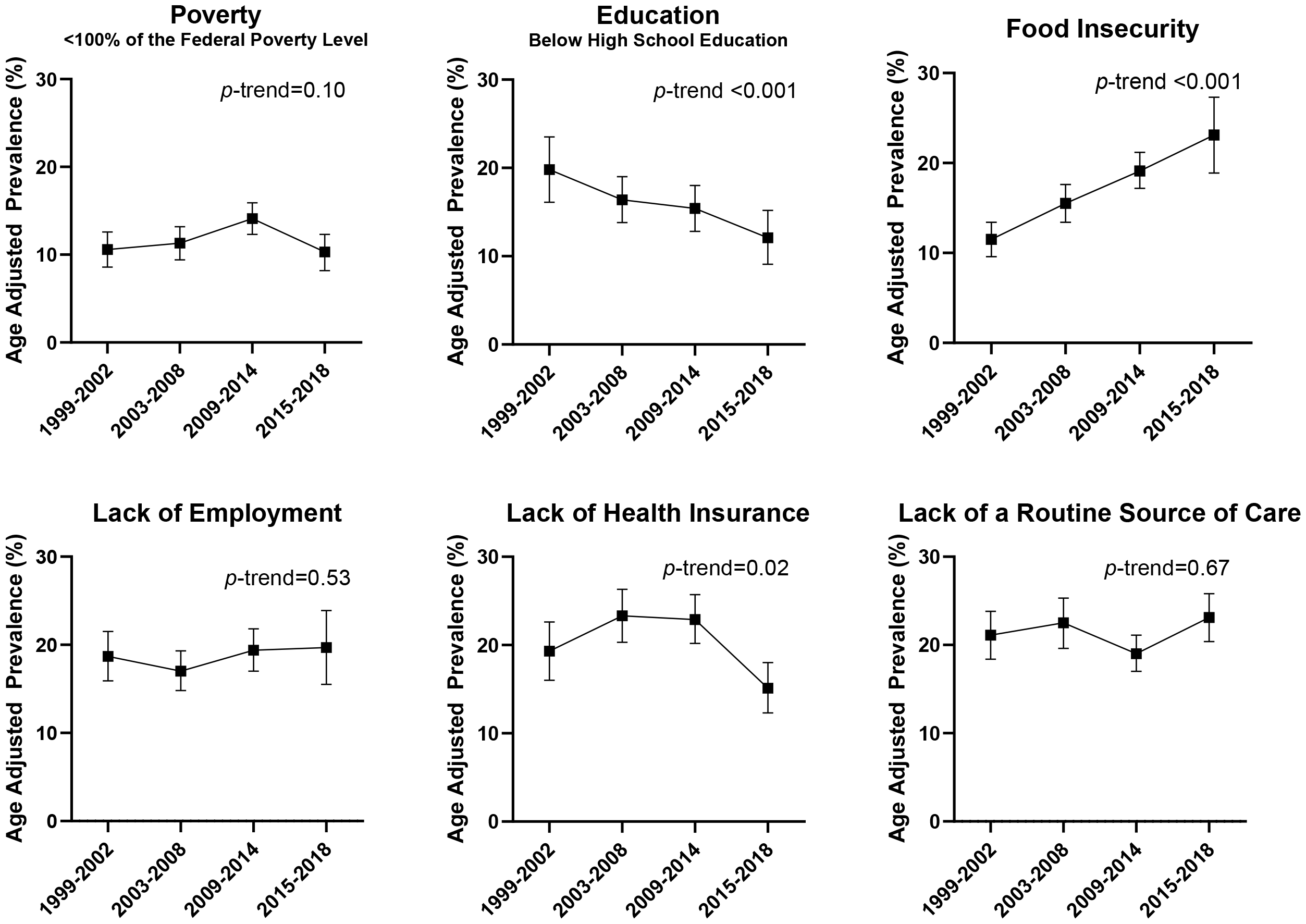
Age-adjusted prevalence of adverse social determinants of health, 1999-2018. Prevalence estimates were age-standardized to the 2010 U.S. Census. *p*-trend was significant for below-high school education, food insecurity, and lack of health insurance.

### Association between Adverse Social Determinants of Health and Mortality

The 15-year adjusted cumulative incidences of mortality and absolute risk differences associated with adverse SDOH are shown in **Figures 2**-**5** and summarized in **Table S7**. The 15-year adjusted cumulative incidences of mortality (ACI) were highest among those with the lowest income (<100% of the federal poverty level; ACI 5.6%, 95% CI 2.8-8.5, **Figure 2**), lowest educational attainment (below high school education; ACI 5.2%, 95% CI 3.2-7.3, **Figure 2**), food insecurity (ACI 4.9%, 95% CI 2.7-7.2, **Figure 3**), and U.S. born participants (ACI 4.0%, 95% CI 2.8-5.2, **Figure 3**). Health insurance and access to care are shown in **Figure 4** and employment status is shown in **Figure 5**. The 15-year absolute risk difference between the lowest and highest income ratio category was 3.2% (95% CI 0.08-6.3). Similarly, the absolute risk difference between a below-high school education and an above-high school education was 2.7% (95% CI 0.4-4.9), and the difference between food insecurity and food security was 2.0% (95% CI -0.04, 4.4). Accumulating a more significant number of social risks was linearly associated with mortality at 15 years (**Figure 5**).

**Figure 2.**
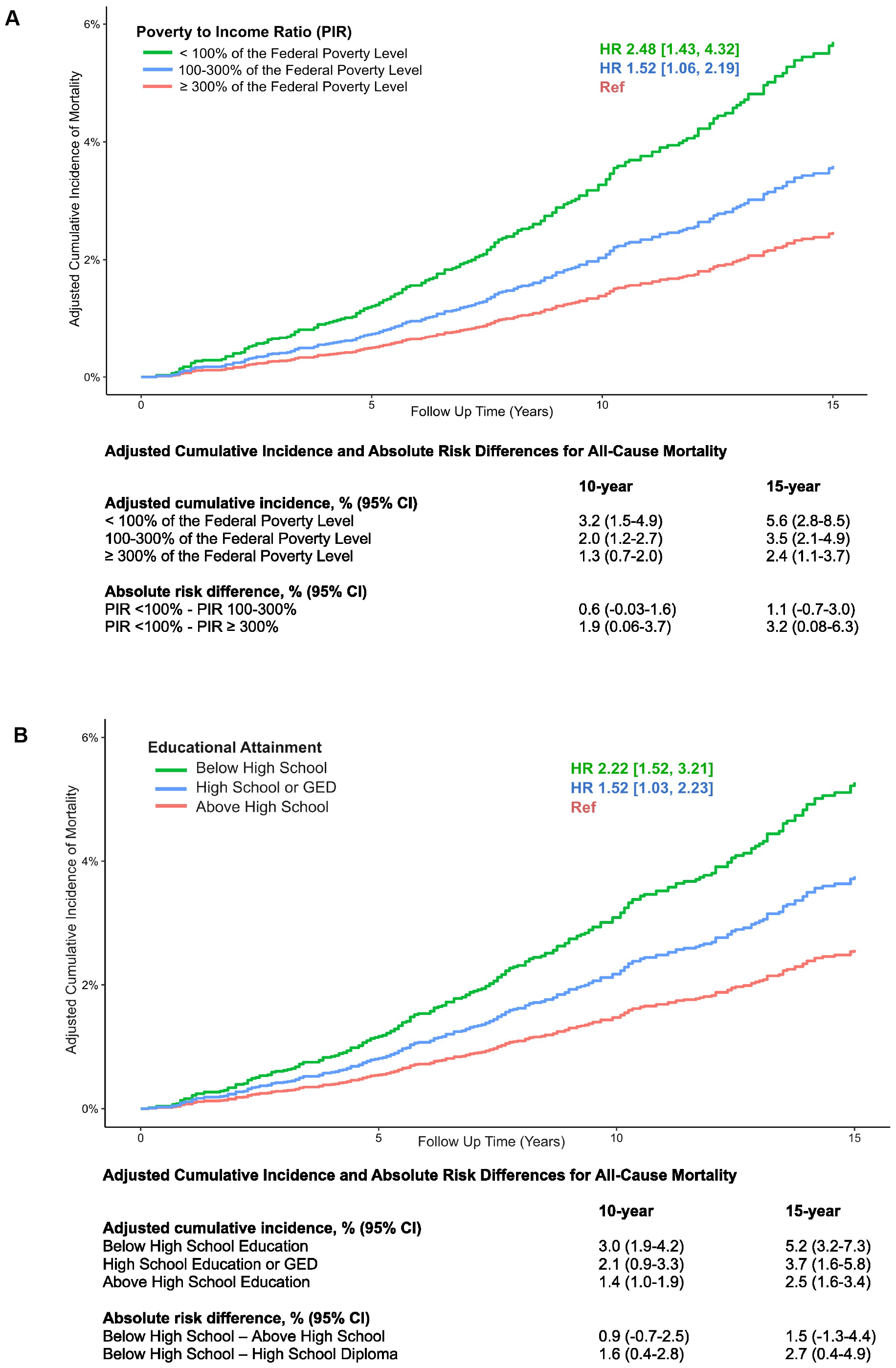
Adjusted cumulative incidences and 10-and 15-year risk differences in adjusted cumulative incidences of all-cause mortality for (A) poverty-to-income ratio and (B) educational attainment. The adjusted cumulative incidences were calculated using confounder-adjusted survival curves employing the G-Formula method for covariate adjustment. Hazard ratios were derived from survey weighted multivariable adjusted Cox regression models.

**Figure 3.**
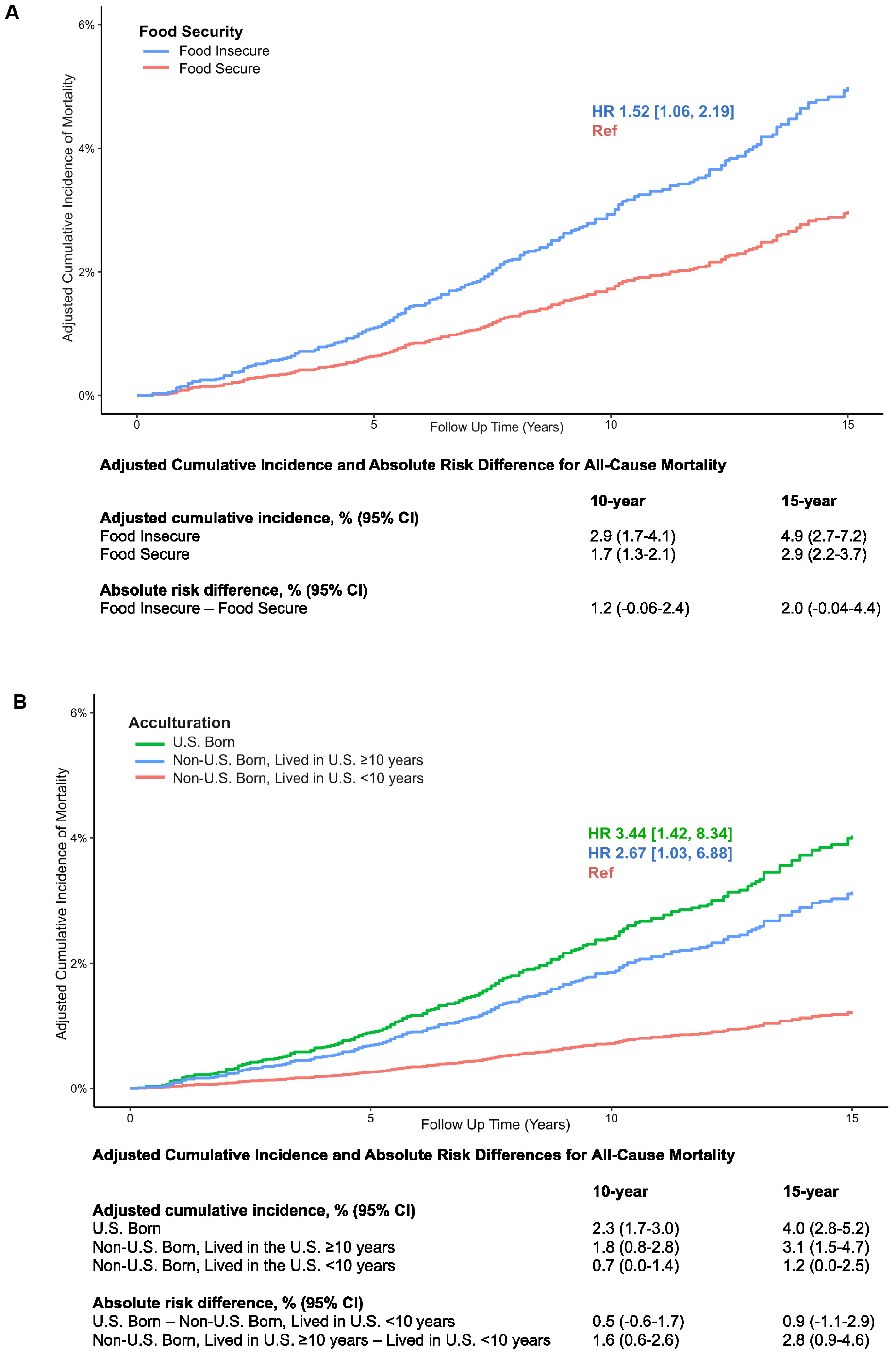
Adjusted cumulative incidences and 10-and 15-year risk differences in adjusted cumulative incidences of all-cause mortality for (A) food security and (B) acculturation. The adjusted cumulative incidences were calculated using confounder-adjusted survival curves employing the G-Formula method for covariate adjustment. Hazard ratios were derived from survey weighted multivariable adjusted Cox regression models.

**Figure 4.**
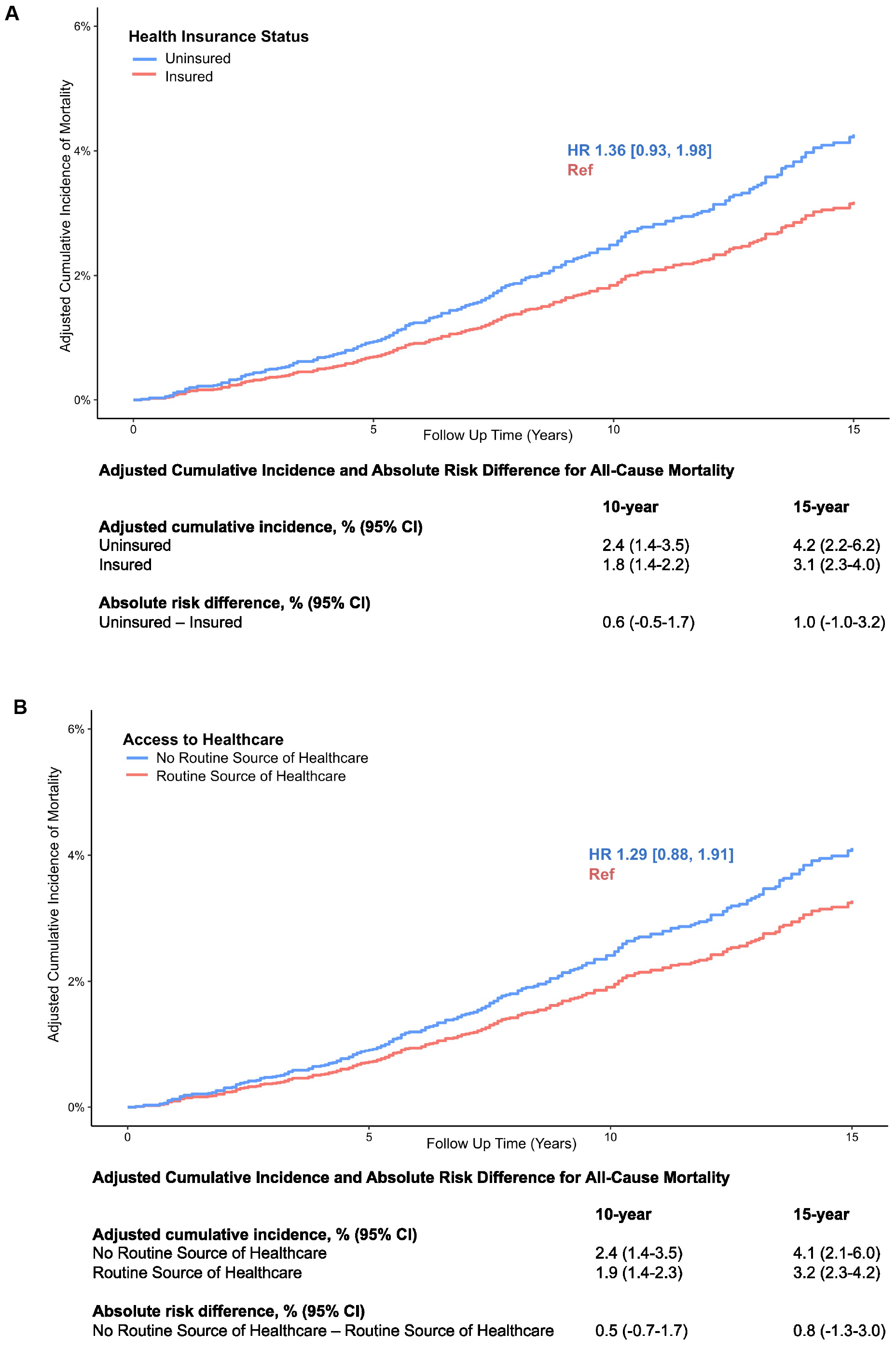
Adjusted cumulative incidences and 10-and 15-year risk differences in adjusted cumulative incidences of all-cause mortality for (A) health insurance status and (B) access to healthcare. The adjusted cumulative incidences were calculated using confounder-adjusted survival curves employing the G-Formula method for covariate adjustment. Hazard ratios were derived from survey weighted multivariable adjusted Cox regression models.

**Figure 5.**
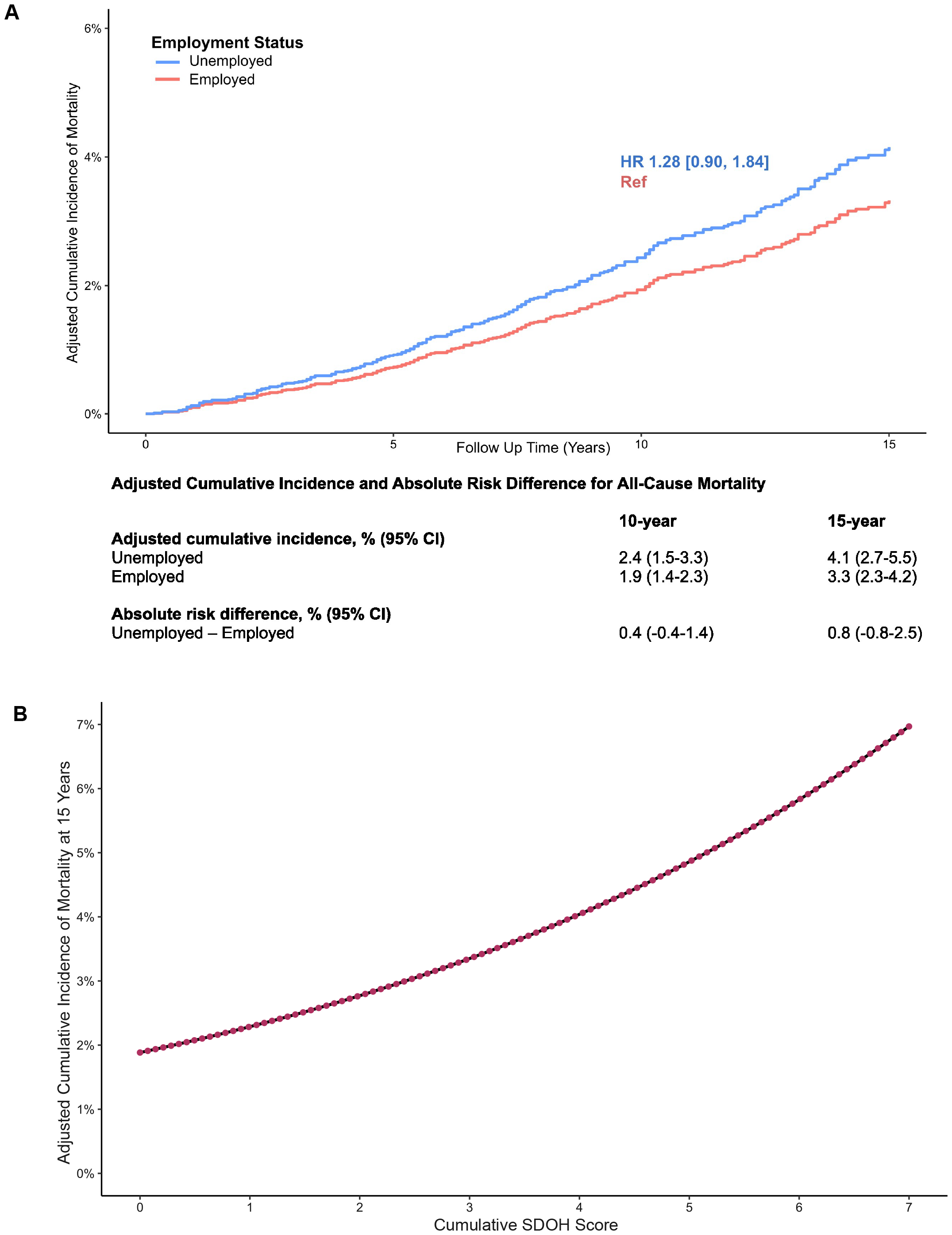
(A) Adjusted cumulative incidences and 10-and 15-year risk differences in adjusted cumulative incidence of all-cause mortality for employment status. (B) 15-year adjusted cumulative incidence of all-cause mortality for increasing cumulative social determinants of health score. The adjusted cumulative incidences were calculated using confounder-adjusted survival curves employing the G-Formula method for covariate adjustment. Hazard ratios were derived from survey weighted multivariable adjusted Cox regression models.

The hazard ratios for mortality associated with each SDOH are shown in **Table S8**. In multivariable-adjusted models, the strongest associations with mortality were observed for nativity/acculturation, income, education, and food security. Compared with recent immigrants, individuals born in the U.S. had 3.44-fold [95% CI 1.42-8.34] higher mortality and immigrants residing in the U.S. for ≥10 years had 2.67-fold [95% CI 1.03-6.88] higher mortality. Compared to those with the highest income (≥300% of the federal poverty level), those with the lowest income (<100% of the federal poverty level) had 2.48-fold [95% CI 1.43-4.32] higher mortality and those with income 100-300% of the federal poverty level had 1.52-fold [95% CI 1.06-2.19] higher mortality. A below-high school education was associated with 2.22-fold [95% CI 1.52-3.21] higher mortality and a high school education was associated with 1.52-fold [95% CI 1.03-2.23] higher mortality than an above-high school education. Food insecurity was associated with 73% [HR 1.73, 95% CI 1.20-2.51] higher mortality. Health insurance, a routine source of healthcare, and employment status were not significantly associated with mortality. Using a cumulative SDOH score, each 1-point increase in adverse SDOH exposure was associated with 33% [HR 1.33, 95% CI 1.16-1.52] higher all-cause mortality.

## Discussion

In this nationally representative sample of community-dwelling adults without major chronic diseases, we demonstrate a pronounced increase in all-cause mortality associated with adverse SDOH. Factors such as poverty, low educational attainment, food insecurity, and acculturation play a more significant role in mortality risk than healthcare access or employment. The compound effects of multiple adverse SDOH can markedly worsen an individual’s health and potentially place them at risk of accumulating additional social risks. In our study, educational attainment improved over the study period, but the sharp increase in food insecurity over the same period is a concerning trend.

Given the breadth of literature on SDOH in disease states and among adolescents,^5,19^ it is perhaps not surprising that a similar relationship exists among apparently healthy adults. However, the magnitude of impact observed is striking. We identified a 5.6% cumulative incidence of mortality due to poverty, while accumulating an increasing number of common social risks linearly increased mortality further in the *absence* of known major chronic disease. These findings have implications for those seeking to impress upon policymakers the importance of SDOH in determining the future health, productivity, and longevity of their healthiest adult constituents.

In the U.S., healthcare-associated expenditures vastly outpace funding for social services, and rising medical costs outpace other high-income countries without conveying improved health outcomes.^1,2^ This ‘American Health Care Paradox’ has been attributed to the unique political and economic history and the social premium on individualism of the U.S., which limits public support for expanded funding for social services.^2^ As a result of such policy decisions in the face of tremendous evidence of the influence of SDOH on health outcomes, many healthcare organizations have moved towards measuring or addressing SDOH once individual patients contact the healthcare system.^20–24^ Other hospital systems strive to reach beyond the exam room and foster deeper community partnerships.^21,25^ While these efforts are regarded as laudable by many, they provoke controversy regarding the responsibility of health systems in directly confronting SDOH.^24,26–28^ Screening and referral to existing public social services, which are woefully underfunded and often unable to provide needed resources, is unlikely to produce population-level shifts in health.^22,23^ At the same time, more innovative, community-centered approaches are only available among select hospital systems.^25^ Considering the existing literature and our findings, point-of-care intervention on individual social needs may miss important opportunities for primary prevention, which more widespread social services could provide. Yet there is not a clear path forward to re-imagining health in the U.S. without support for reshaping public policy and the social contract in the U.S.^29,30^

To our knowledge, this is the first report of the impact of adverse SDOH on mortality within relatively healthy adults without known major chronic diseases. This analysis has several strengths, including the use of objectively determined chronic disease states, a nationally representative population, and a long duration of follow-up. These analyses would be further improved by including incident chronic diseases as outcomes, which are unavailable in NHANES. Similarly, we were unable to include variables related to social cohesion, structural racism, income inequality, or neighborhood and community environments, which are significant SDOH.^31–33^

In conclusion, our findings challenge the conventional healthcare-centric approach to preventative care, emphasizing the need for proactive public health interventions targeting SDOH.^34^ Identifying and intervening on SDOH once a patient has developed a disease may overlook years of accumulated risk. Our findings strengthen the argument for societal intervention on SDOH. We can significantly reduce population-wide morbidity and mortality by addressing social determinants early.

## Data Availability

All data are publicly available at https://wwwn.cdc.gov/nchs/nhanes/default.aspx

## Disclosures

The authors have no conflicts of interest to disclose.

## Funding

The research reported in this publication was supported by the National Heart, Lung, and Blood Institute of the National Institutes of Health under Award Number R38HL143584.

## Notes

### Competing Interest Statement

The authors have declared no competing interest.

### Author Declarations

The data used in this study are publicly available at https://wwwn.cdc.gov/nchs/nhanes/default.aspx.

