## Supplemental Tables for "sSocial Determinants of Health and Cumulative Incidence of Mortality among U.S. Adults without Major Chronic Diseases"

| Table S1. Exclusion criteria and variable definitions | |
|---|---|
| **Variable** | **Definition** |
| Hypertension | Systolic blood pressure ≥130 mmHg OR diastolic blood pressure ≥80 mmHg OR current use of an antihypertensive medication |
| Diabetes | Hemoglobin A1c (HbA1c) ≥6.5% (44.0 mmol/mol) OR current use of insulin OR current use of an anti-hyperglycemic medication |
| Class III Obesity | Body mass index (BMI) ≥40 $kg/m^2$ |
| Hyperlipidemia | Total cholesterol ≥240 mg/dL (6.5 mmol/L) OR current use of a lipid-lowering medication |
| Chronic kidney disease | Estimated glomerular filtration rate <60 ml/min per $1.73m^2$ or urinary albumin-to-creatinine ratio ≥30 mg/g (3 mmol/mol) |
| History of cardiovascular disease | Ever been told by a medical professional that you have had coronary heart disease, angina pectoris, heart failure, a stroke, or a heart attack. |
| History of cancer | Ever been told by a medical professional that you have had a cancer or malignancy of any kind. |
| History of respiratory disease | Ever been told by a medical profession that you have chronic bronchitis, emphysema, asthma, or chronic obstructive pulmonary disease. |
| History of liver disease | Ever been told by a medical professional that you have had any kind of liver condition. |
| History of arthritis | Ever been told by a medical professional that you have arthritis. |
| History of dialysis use | Underwent dialysis in the last 12 months. |
| Hepatitis B | Positive hepatitis B core antibody with duplicate testing. |
| Hepatitis C | Positive hepatitis C core antibody with duplicate testing. |
| Human immunodeficiency virus (HIV) | Positive HIV-1 or HIV-2 antibody with duplicate testing and/or confirmatory testing. |
| Pregnancy | Positive urine pregnancy test. |

| **Table S2. Variable Definitions for Social Determinants of Health** | | | |
|---|---|---|---|
| **Variable** | **Domain†** | **NHANES Question‡ / Variable Definition** | **Variable Levels** |
| Poverty to Income Ratio | Economic Stability | Total family income was reported by participants as a range of values (in dollars). NHANES then calculated a family poverty-to-income ratio. | <100%, 100-300%, and ≥300% of the federal poverty level. |
| Educational Attainment | Education Access and Quality | What is the highest grade or level of school you have completed, or the highest degree you have received? | Below high school, high school or general education diploma, and above high school. |
| Food Insecurity | Economic Stability | Participants completed an 18-item U.S. Household Food Security Survey Module. | Food secure (zero affirmative responses) and food insecure (≥1 positive response). |
| Employment | Economic Stability | What kind of work did you do in the last week? | Employed (currently working, student, or retired) and unemployed (not currently working). |
| Health Insurance | Health Care Access and Quality | Are you covered by health insurance or some other kind of health care plan? | Insured and uninsured. |
| Routine source of care | Health Care Access and Quality | Is there a place that you usually go when you are sick or need advice about your health? What kind of place do you go most often? | Routine source of care (there is at least one source of care that is not the emergency room) and lack of a routine source of care (no source of care or only rely on the emergency room). |
| Acculturation | Social and Community Context | What country were you born in? If you were not born in the U.S., how long have you been in the U.S. | U.S. born, non-U.S. born and lived in the U.S. for ≥10 years, and non-U.S. born and lived in the U.S. <10 years. |
| Cumulative SDOH score | All of the above | One point value was assigned to each adverse social determinant: PIR <100%, below high school education, food insecurity, unemployment, lack of health insurance, lack of a routine source of healthcare, born in the U.S. | A cumulative score was obtained by summing the number of points (range 0-7). |

† As defined by Healthy People 2030. Available at https://health.gov/healthypeople/priority-areas/social-determinants-health
‡ Details available at https://wwwn.cdc.gov/nchs/nhanes/default.aspx

| Table S3. Missingness prior to imputation | |
| --- | --- |
| | **N (%)** |
| Smoking status | 12 (0.1) |
| Hemoglobin A1c | 19 (0.2) |
| Body mass index | 105 (1.0) |
| Systolic blood pressure | 541 (4.9) |
| Total cholesterol | 876 (8.0) |
| Serum creatinine | 900 (8.2) |

| Table S4. Participant Characteristics Prior to Imputation (N=10,320). | | | | | |
|---|---|---|---|---|---|
| | **Overall** | **Non-Hispanic White** | **Non-Hispanic Black** | **Hispanic†** | **Other** |
| N | 10,320 | 4,116 | 1,698 | 3,433 | 1,073 |
| Age | 35.0 (11.2) | 36.2 (11.6) | 32.2 (10.0) | 32.9 (9.9) | 33.6 (9.8) |
| Female sex | 52.0 | 51.9 | 52.6 | 50.1 | 56.1 |
| Race or ethnicity | | | | | |
| Non-Hispanic White | 64.5 | 100 | 0 | 0 | 0 |
| Mexican American | 12.2 | 0 | 0 | 62.6 | 0 |
| Other Hispanic | 7.3 | 0 | 0 | 37.4 | 0 |
| Non-Hispanic Black | 9.1 | 0 | 100 | 0 | 0 |
| Other Race | 6.9 | 0 | 0 | 0 | 100 |
| Education | | | | | |
| Below High School | 15.0 | 7.7 | .17.8 | 39.5 | 9.3 |
| High School/GED | 21.8 | 21.9 | 26.0 | 22.0 | 14.8 |
| Above High School | 63.3 | 70.4 | 56.2 | 38.5 | 75.9 |
| Poverty-to-Income Ratio | | | | | |
| <100% FPL | 14.2 | 8.5 | 24.3 | 29.1 | 15.1 |
| 100-300% FPL | 35.7 | 31.8 | 44.3 | 44.3 | 30.3 |
| ≥300 FPL | 50.1 | 59.7 | 31.3 | 23.6 | 54.5 |
| Food insecurity | 22.3 | 14.4 | 36.7 | 43.5 | 18.0 |
| Uninsured | 25.0 | 17.3 | 32.5 | 49.1 | 19.3 |
| Lack of employment | 19.2 | 17.6 | 21.8 | 22.7 | 20.9 |
| Lack a regular source of care | 26.1 | 21.2 | 28.2 | 40.5 | 28.7 |
| Acculturation | | | | | |
| US Born | 82.0 | 94.8 | 89.0 | 43.8 | 39.1 |
| US Born, Lived in US ≥10 years | 10.7 | 2.9 | 6.6 | 35.6 | 31.9 |
| Non-US Born, Lived in US <10 years | 7.4 | 2.4 | 4.5 | 20.6 | 29.0 |
| Cumulative SDOH score | 2.5 (1.1) | 2.4 (1.0) | 2.9 (1.3) | 2.7 (1.3) | 1.9 (1.2) |
| Smoking status | | | | | |
| Never | 61.2 | 57.1 | 68.0 | 67.8 | 72.0 |
| Former | 16.5 | 18.6 | 6.9 | 14.9 | 13.0 |
| Current | 22.3 | 24.3 | 25.1 | 17.2 | 15.0 |
| Heavy alcohol use | 15.3 | 14.9 | 9.9 | 21.8 | 7.7 |
| Intravenous drug use | 6.8 | 8.2 | 3.4 | 5.1 | 2.8 |
| Body Mass Index, kg/m$^2$ | 26.2 (4.8) | 25.9 (4.7) | 27.2 (5.3) | 27.2 (4.6) | 24.8 (4.6) |
| Hemoglobin A1c, % | 5.2 (0.3) | 5.2 (0.3) | 5.3 (0.4) | 5.3 (0.3) | 5.3 (0.3) |
| Total Cholesterol, mg/dL | 181.8 (29.5) | 182.8 (29.4) | 176.3 (30.5) | 181.5 (29.5) | 180.4 (28.8) |

| | | | | | |
|---|---|---|---|---|---|
| Systolic Blood Pressure, mmHg | 111.5 (8.8) | 111.6 (9.1) | 112.9 (8.6) | 111.3 (8.8) | 109.9 (9.2) |
| eGFR, ml/min per 1.73m$^2$ | 106.6 (15.9) | 104.1 (15.6) | 102.3 (16.8) | 114.7 (13.8) | 111.6 (13.8) |

† Hispanic includes both Mexican American and Other Hispanic.

Abbreviations: eGFR, estimated glomerular filtration rate; FPL, federal poverty level; GED, general education diploma.

| Table S5. Participant Characteristics (N=11,413). | | | | | |
|---|---|---|---|---|---|
| | **Overall** | **Non-Hispanic White** | **Non-Hispanic Black** | **Hispanic†** | **Other** |
| N | 11,413 | 4,464 | 1,967 | 3,752 | 1,230 |
| Age | 34.9 (11.2) | 36.1 (11.7) | 32.4 (10.1) | 32.9 (9.8) | 33.8 (10.0) |
| Female sex | 51.9 | 52.0 | 51.9 | 50.0 | 55.9 |
| Race or ethnicity | | | | | |
| Non-Hispanic White | 63.7 | 100 | 0 | 0 | 0 |
| Mexican American | 12.2 | 0 | 0 | 62.6 | 0 |
| Other Hispanic | 7.3 | 0 | 0 | 37.4 | 0 |
| Non-Hispanic Black | 9.6 | 0 | 100 | 0 | 0 |
| Other Race | 7.2 | 0 | 0 | 0 | 100 |
| Education | | | | | |
| Below High School | 15.4 | 7.9 | 18.0 | 40.8 | 9.9 |
| High School/GED | 21.9 | 22.1 | 26.4 | 21.7 | 14.3 |
| Above High School | 62.7 | 70.0 | 55.6 | 37.5 | 75.9 |
| Poverty-to-Income Ratio | | | | | |
| <100% FPL | 14.8 | 9.0 | 24.6 | 29.7 | 15.9 |
| 100-300% FPL | 35.7 | 31.8 | 43.8 | 47.2 | 31.2 |
| ≥300 FPL | 49.5 | 59.3 | 31.6 | 23.1 | 52.9 |
| Food insecurity | 22.4 | 14.5 | 36.0 | 43.6 | 18.1 |
| Uninsured | 25.3 | 17.5 | 32.2 | 49.9 | 19.1 |
| Lack of employment | 19.6 | 18.0 | 22.8 | 22.6 | 21.5 |
| Lack a regular source of care | 26.4 | 21.5 | 28.2 | 41.0 | 28.4 |
| Acculturation | | | | | |
| US Born | 81.4 | 94.7 | 88.2 | 43.3 | 37.6 |
| US Born, Lived in US ≥10 years | 10.9 | 3.0 | 6.8 | 35.9 | 32.7 |
| Non-US Born, Lived in US <10 years | 7.7 | 2.4 | 5.0 | 20.8 | 29.7 |
| Cumulative SDOH score | 2.5 (1.1) | 2.4 (1.0) | 2.9 (1.3) | 2.8 (1.3) | 1.9 (1.2) |
| Smoking status | | | | | |
| Never | 61.6 | 57.4 | 68.5 | 68.0 | 72.2 |
| Former | 16.0 | 18.1 | 6.6 | 14.7 | 12.8 |
| Current | 22.4 | 24.5 | 24.9 | 17.3 | 15.0 |
| Heavy alcohol use | 20.9 | 20.0 | 17.3 | 27.7 | 16.4 |
| Intravenous drug use | 6.5 | 8.0 | 3.1 | 5.0 | 2.5 |
| Body Mass Index, $kg/m^2$ | 26.2 (4.8) | 25.9 (4.8) | 27.2 (5.3) | 27.2 (4.6) | 24.8 (4.6) |
| Hemoglobin A1c, % | 5.2 (0.3) | 5.2 (0.3) | 5.3 (0.4) | 5.3 (0.3) | 5.3 (0.3) |
| Total Cholesterol, mg/dL | 181.8 (29.6) | 182.8 (29.4) | 177.0 (30.4) | 181.6 (29.7) | 180.7 (29.0) |

| | | | | | |
|---|---|---|---|---|---|
| Systolic Blood Pressure, mmHg | 111.6 (9.1) | 111.7 (9.1) | 113.0 (8.8) | 111.3 (8.8) | 110.3 (9.2) |
| eGFR, ml/min per 1.73m$^2$ | 106.4 (16.2) | 104.1 (15.8) | 102.5 (17.0) | 114.4 (14.2) | 110.8 (14.8) |

† Hispanic includes both Mexican American and Other Hispanic.

Abbreviations: eGFR, estimated glomerular filtration rate; FPL, federal poverty level; GED, general education diploma; SDOH, social determinants of health.

| Table S6. Trends in Age-Adjusted* Prevalence of Social Determinants of Health, 1999-2018. | | | | | | |
|---|---|---|---|---|---|---|
| | **Overall** | **1999-2002** | **2003-2008** | **2009-2014** | **2015-2018** | ***p*-trend** |
| Poverty-to-Income Ratio | | | | | | |
| <100% FPL | 11.8 (10.8, 12.8) | 10.6 (8.6, 12.6) | 11.3 (9.4, 13.2) | 14.1 (12.3, 15.9) | 10.3 (8.2, 12.3) | 0.10 |
| 100-300% FPL | 33.2 (31.1, 35.3) | 34.4 (29.4, 39.4) | 38.1 (34.9, 41.2) | 30.6 (26.4, 34.8) | 32.5 (28.4, 36.6) | 0.91 |
| ≥300% FPL | 55.1 (52.7, 57.4) | 55.0 (49.4, 60.5) | 50.6 (47.1, 54.1) | 55.3 (50.6, 60.0) | 57.2 (52.5, 61.9) | 0.46 |
| Educational Attainment | | | | | | |
| Below high school | 15.5 (14.1, 17.0) | 19.8 (16.1, 23.5) | 16.4 (13.8, 19.0) | 15.4 (12.8, 18.0) | 12.1 (9.1, 15.2) | <0.001 |
| High school/GED | 19.9 (18.1, 21.6) | 20.3 (15.5, 25.1) | 22.4 (19.7, 25.1) | 17.4 (14.8, 20.0) | 20.3 (15.5, 25.1) | 0.50 |
| Above high school | 64.5 (62.2, 66.8) | 60.0 (55.2, 64.7) | 61.1 (57.9, 64.3) | 67.2 (63.7, 70.7) | 67.5 (61.1, 74.0) | 0.02 |
| Food Insecure | 17.5 (16.2, 18.9) | 11.5 (9.6, 13.4) | 15.5 (13.4, 17.6) | 19.1 (17.2, 21.2) | 23.1 (18.9, 27.3) | <0.001 |
| Unemployed | 18.9 (17.3, 20.5) | 18.7 (15.9, 21.5) | 17.0 (14.8, 19.3) | 19.4 (17.0, 21.8) | 19.7 (15.5, 23.9) | 0.53 |
| Uninsured | 20.5 (19.0, 21.9) | 19.3 (16.0, 22.6) | 23.3 (20.3, 26.3) | 22.9 (20.2, 25.7) | 15.1 (12.3, 18.0) | 0.02 |
| No usual source of care | 21.2 (19.9, 22.5) | 21.1 (18.4, 23.8) | 22.5 (19.6, 25.3) | 19.0 (17.0, 21.1) | 23.1 (20.4, 25.8) | 0.67 |

* Age-standardized to the 2010 U.S. Census
Abbreviations: FPL, federal poverty level; GED, general education diploma.

| Table S7. Adjusted cumulative incidence of mortality and absolute risk difference by social determinant of health | | | | |
|---|---|---|---|---|
| | **10-year adjusted cumulative incidence (95% CI)** | **15-year adjusted cumulative incidence (95% CI)** | **10-year risk difference (95% CI)** | **15-year risk difference (95% CI)** |
| **Poverty-to-Income Ratio (PIR)** | | | | |
| <100% FPL | 3.2% (1.5-4.9) | 5.6% (2.8-8.5) | | |
| 100-300% FPL | 2.0% (1.2-2.7) | 3.5% (2.1-4.9) | 0.6% (-0.03-1.6) [a] | 1.1% (-0.7-3.0) [a] |
| ≥300% FPL | 1.3% (0.7-2.0) | 2.4% (1.1-3.7) | 1.9% (0.06-3.7) [b] | 3.2% (0.08-6.3) [b] |
| **Educational Attainment** | | | | |
| Below high school | 3.0% (1.9-4.2) | 5.2% (3.2-7.3) | | |
| High school/GED | 2.1% (0.9-3.3) | 3.7% (1.6-5.8) | 0.9% (-0.7-2.5) [c] | 1.5% (-1.3-4.4) [c] |
| Above high school | 1.4% (1.0-1.9) | 2.5% (1.6-3.4) | 1.6% (0.4-2.8) [d] | 2.7% (0.4-4.9) [d] |
| **Employment Status** | | | | |
| Unemployed | 2.4% (1.5-3.3) | 4.1% (2.7-5.5) | 0.4% (-0.4-1.4) | 0.8% (-0.8-2.5) |
| Employed | 1.9% (1.4-2.3) | 3.3% (2.3-4.2) | | |
| **Food Security** | | | | |
| Food insecure | 2.9% (1.7-4.1) | 4.9% (2.7-7.2) | 1.2% (-0.06-2.4) | 2.0% (-0.04-4.4) |
| Food secure | 1.7% (1.3-2.1) | 2.9% (2.2-3.7) | | |
| **Health Insurance** | | | | |
| Uninsured | 2.4% (1.4-3.5) | 4.2% (2.2-6.2) | 0.6% (-0.5-1.7) | 1.0% (-1.0-3.2) |
| Insured | 1.8% (1.4-2.2) | 3.1% (2.3-4.0) | | |
| **Access to Care** | | | | |
| Lack of a routine source of healthcare | 2.4% (1.2-3.5) | 4.1% (2.1-6.0) | 0.5% (-0.7-1.7) | 0.8% (-1.3-3.0) |
| Routine source of healthcare | 1.9% (1.4-2.3) | 3.2% (2.3-4.2) | | |
| **Nativity and Acculturation** | | | | |
| U.S. Born | 2.3% (1.7-3.0) | 4.0% (2.8-5.2) | | |
| Non-U.S. Born, Lived in U.S. ≥ 10 years | 1.8% (0.8-2.8) | 3.1% (1.5-4.7) | 0.5% (-0.6-1.7) [e] | 0.9% (-1.1-2.9) [e] |
| Non-U.S. Born, Lived in U.S. < 10 years | 0.7% (0.0-1.4) | 1.2% (0.0-2.5) | 1.6% (0.6-2.6) [f] | 2.8% (0.9-4.6) [f] |

[a] Risk difference represents: PIR <100% – PIR 100-300%
[b] Risk difference represents: PIR <100% – PIR >300%
[c] Risk difference represents: below high school – above high school
[d] Risk difference represents: below high school – high school
[e] Risk difference represents: U.S. Born – Non-U.S. Born, Lived in the U.S. ≥ 10 years
[f] Risk difference represents: U.S. Born – Non-U.S. Born, Lived in the U.S. < 10 years

Models adjusted for age, sex, race or ethnicity, smoking status, body mass index, hemoglobin A1c, total cholesterol, systolic blood pressure, eGFR, survey year, alcohol use, and injection drug use.

Abbreviations: CI, confidence interval; FPL, federal poverty level; GED, general education diploma; PIR, poverty to income ratio; U.S., United States.

| Table S8. Associations between social determinants of health and mortality. | | | | | |
|---|---|---|---|---|---|
| | N [†] | Crude [a] | Model 1 [b] | Model 2 [c] | Model 3 [d] |
| | | HR [95% CI] | HR [95% CI] | HR [95% CI] | HR [95% CI] |
| Poverty-to-Income Ratio | 10,433 | | | | |
| <100% FPL | | 1.64 [1.02, 2.62] | 3.12 [1.86, 5.24] | 2.40 [1.38, 4.18] | 2.48 [1.43, 4.32] |
| 100-300% FPL | | 1.19 [0.82, 1.71] | 1.72 [1.19, 2.49] | 1.49 [1.04, 2.14] | 1.52 [1.06, 2.19] |
| ≥300% FPL | | Referent | Referent | Referent | Referent |
| Education | 11,402 | | | | |
| Below high school | | 2.23 [1.60, 3.11] | 2.72 [1.84, 4.01] | 2.17 [1.49, 3.16] | 2.22 [1.52, 3.21] |
| High school/GED | | 1.54 [1.04, 2.30] | 1.73 [1.18, 2.54] | 1.52 [1.05, 2.21] | 1.52 [1.03, 2.23] |
| Above high school | | Referent | Referent | Referent | Referent |
| Lack of employment | 11,407 | 1.07 [0.75, 1.52] | 1.34 [0.94, 1.93] | 1.28 [0.89, 1.83] | 1.28 [0.90, 1.84] |
| Food insecurity | 11,138 | 1.32 [0.95, 1.83] | 2.10 [1.47, 3.02] | 1.73 [1.20, 2.49] | 1.73 [1.20, 2.51] |
| Nativity and Acculturation | 10,394 | | | | |
| U.S. Born | | 5.10 [2.24, 11.60] | 3.75 [1.56, 9.06] | 3.53 [1.46, 8.52] | 3.44 [1.42, 8.34] |
| U.S. Born, Lived in U.S. ≥10 years | | 4.66 [1.82, 11.96] | 2.70 [1.04, 6.99] | 2.68 [1.04, 6.89] | 2.67 [1.03, 6.88] |
| Non-US Born, Lived in U.S. <10 years | | Referent | Referent | Referent | Referent |
| Lack of health insurance | 11,359 | 1.07 [0.77, 1.51] | 1.67 [1.17, 2.37] | 1.37 [0.94, 1.99] | 1.36 [0.93, 1.98] |
| Lack of regular source of care | 11,413 | 1.08 [0.74, 1.58] | 1.47 [0.99, 2.18] | 1.31 [0.88, 1.94] | 1.29 [0.88, 1.91] |
| Cumulative SDOH score | 11,413 | 1.25 [1.12, 1.41] | 1.45 [1.27, 1.64] | 1.32 [1.15, 1.52] | 1.33 [1.16, 1.52] |

[†] Sample size varies due to variable participant non-response to social determinants of health questions.
[a] Crude
[b] Model 1: adjusted for age, sex, race or ethnicity, and survey year.
[c] Model 2: adjusted for Model 1 + smoking status, body mass index, hemoglobin A1c, total cholesterol, systolic blood pressure, eGFR.
[d] Model 3: adjusted for Model 2 + alcohol use, injection drug use.

Abbreviations: eGFR, estimated glomerular filtration rate; FPL, federal poverty level; GED, general education diploma; HR, hazard ratio; SDOH, social determinants of health; U.S., United States.
